# Association between wasting and feeding practices consumption during 24 hours recall in young children aged 6-23 months in Cambodia: Analysis of Cambodia Demographic and Health Survey 2014

**DOI:** 10.1101/2022.10.24.22281479

**Authors:** Samnang Um, Mom Luon, Yom An, Bunkea Tol

## Abstract

**INTRODUCTION:** World Health Organization (WHO), defines wasting as a child’s weight-for-height z-score (WHZ) below minus 2 SD of the Child’s Growth Standards. In Cambodia, the prevalence of wasting among children under five years old increased from 8% in 2005 to 10% in 2014. The WHO divided food into seven categories, including staple foods, legumes, milk, meat, eggs, fruits, and vegetables. It was advised to consume at least four of these categories daily. According to data from the 2014 Cambodia Demographic and Health Survey (CDHS), 48% of kids between the ages of six and 23 months consumed at least four food groups. We aimed to identify the association between feeding practices consumption and wasting in Cambodian children aged 6-23 months.

**METHODS:** We used existing children’s data from CDHS 2014, two-stage stratified cluster sampling approach to select samples. Data analysis was done by using STATA V16 Survey weights were applied to account for the complex survey design of the CDHS. Descriptive statistics were estimated for key children characteristics, maternal, and household characteristics, geographical regions, and feeding practice consumption. We ran bivariate and multiple logistic regressions to assess the association between wasting and feeding practices consumption in children aged 6-23 months.

**RESULTS:** A total of 1,415 children aged 6–23 months were eligible for the study. This study did not find an association between wasting and feeding practices consumption. In contrast, children aged 9–11 months had 2.3 times of wasting [AOR = 2.3; 95% CI = 1.0–5.0] compared to children aged 6–8 months. When compared to mothers aged 15–19 years, children born from mothers aged 20–34 years and 35–49 years were protected from 60% of wasting [AOR = 0.3; 95% CI = 0.2–0.6] and [AOR = 0.3; 95% CI = 0.1–0.7], respectively. When compared to children from better-off households, children from the poorest and poorer wealthiest households wasted twice as much time [AOR = 1.9; 95% CI 1.1-3.5].

**CONCLUSION:** These findings indicate that feeding practices consumption is not significantly associated with wasting in young children aged 6–23 months in Cambodia. It was found in other studies. Children aged 9–11 months and children from the poorest households were the main predictors of child mortality. However, mothers aged 20–49 years old had decreased odds of wasting. Therefore, it is recommended that interventions and policymakers prioritize promoting diverse food consumption among children in Cambodia. Furthermore, interventions to reduce wasting among children aged 6 to 11 months, children from the lowest household quintile, and children of young mothers should be prioritized.

## INTRODUCTION

In 2020, approximately 46 million (7%) of children aged 0 to59 months are wasting away, with 14 million (2%) severely [1]. When a child’s weight-for-height z-score (WHZ) is below minus 2SD of the Child’s Growth Standards, it is deemed to be wasting by WHO [2]. Whereas 14.6% of them are wasting lives in South Asia [1]. Undernutrition in developing nations has been linked to over, 45% of deaths in children under five (U5) [3]. U5 wasting rates increased from 8% in 2005 to 10% in 2014, according to the most recent Cambodia Demographic and Health Survey (CDHS), [4]. The percentage of child wasting varied from10% in rural areas to 8% in urban areas and was observed at 15% in provinces of Takeo, and Oddar Meanchey, respectively [4]. Additionally, it is estimated that each child will cost USD $ 113 in specialized medical care, including therapeutic feeding [5]. Using a questionnaire and data from interviews conducted within the previous day, the consumption of child feeding practices was assessed at the household or individual level [6]. The WHO divided food into seven categories, namely staple foods, legumes, milk, meat, eggs, fruits, and vegetables. It was advised to consume at least four these categories each day [7]. In 2014, 48% of children in Cambodia aged 6-23 months reported eating from four or more food groups [4]. Additionally, compared to children who are healthy, children who are wasted have a higher risk of dying [8]. Southeast Asia’s Cambodia is an agricultural country. With total population of 16 million as of 2019, it borders Thailand to the west, Laos and Thailand to the north, the Gulf of Thailand to the southwest, and Vietnam to the east and the south, [9]. One of the Sustainable Development Goals (SDG) of the United Nations is to end world hunger by 2030. Achieving food security, enhancing nutrition, and promoting sustainable agriculture are key milestones toward this goal [9, 10]. Incorporating government initiatives aimed at promoting infant and maternal care, as well as expanding nutrition and child health programs, are just a few of the numerous advancements that Cambodia had made [11]. Malnutrition is made more likely by poor infant and young child feeding (IYCF) practices [12]. In developing countries, the custom of offering complimentary food diversity is based on the local geography and is more likely to be tailored to the social conditions of the family. The majority of the food consumed by underprivileged families had to no nutritional value [13]. In comparison to people who live in rural settings, those who live in urban areas are more likely to have a higher incomes and consumed a wider variety foods [14]. Children born from mothers with low body mass index (<18.5 kg/m^2^) are wasted in all six South Asia countries between 2009 to 2016. This research focused on the factors that cause wasting in young children age under five [15]. Additionally, a child born from late birth order, to man, whose mother reported a limited level of education, was short height, lacked access to improved water source, and live in a poor household was also at risk of wasting in various countries [15]. Despite these, factors such as poor household wealth index, inadequate sanitation and drinking water [16, 17], child age, child sex, older maternal age, mother without formal education, residential area are known increased risk of wasting among children of 6-23 months [18]. Children of 6-23□months who received minimally diversified diet, frequent meals, and acceptable diet were associated had lower rate of undermatron [17, 18]. A lack of dietary variety was found to be risk of weight loss in serval studied [19]. Despite the literature’s availability, there hasn’t been study using CDHS data to examine the relationship between wasting and dietary diversity consumption among children aged 6-23 months in Cambodia. Therefore, we sought to investigate the child feeding practices consumption and socio-demographic factors associated with wasting in young children in Cambodia aged 6-23 months.

## MATERIALS AND METHODS

### Data

We used existing children’s data from Cambodia Demographic and Health Survey (CDHS) 2014. The CDHS is a nationally representative population-based household survey that is regularly conducted roughly every 5 years. Data collection procedures have been described and published in the CDHS 2014 report [4]. The survey employed a two-stage stratified cluster sampling to collect the samples from all 19 sampling domains, which were further divided into 38 sampling strata between urban and rural. In the first stage, 611 enumeration areas (EAs) number of households residing in the enumerator’s area or clusters (188 urban EAs and 423 rural EAs) were selected from 4,245 urban EAs and 24,210 rural EAs respectively using probability proportional to EA size. The second stage, a fixed number of households were selected from each cluster (24 urban and 28 rural) through systematic sampling [4]. We restricted analysis to only data of children aged between 6-23 months, living with their mothers or caregivers, and children with valid anthropometric measurements of height and weight were included. Finally selected for further analysis (**Fig 1**).

**Figure 1.**
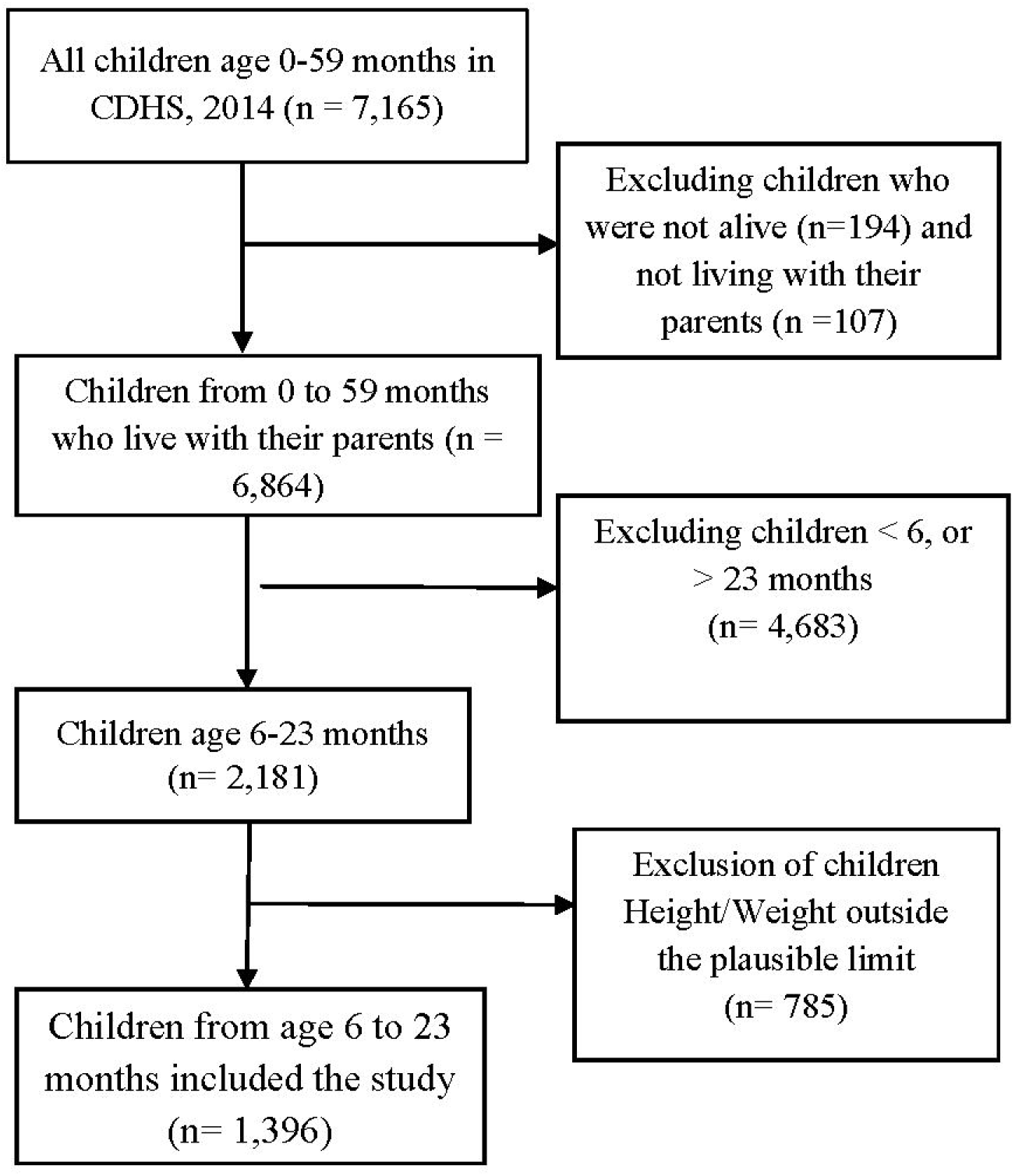
Sample selection diagram

### Measurements Outcome variable

Wasting is the outcome variable used in the study. For children in CDHS, anthropometric measurements of height and weight were taken one to three times, and age was determined using the child’s reported age or date of birth. All weights were measured using a digital SECA scale created with UNICEF’s approval, and all heights were measured with Shorr Productions measuring boards. All infants younger than 24 months had their recumbent length measured, and all children older than 24 months had their standing height was measured [4]. Then, the original variable was then converted into the dichotomous variable, with WHZ-score <u>be low</u> minus 2 SD denoting **wasting** =1, and WHZ-score greater than minus 2 SD denoting **non wasting**=0, respectively.

### Explanatory variables

#### Minimum Dietary diversity (MDD)

We used the Infant and Young Children Feeding guidelines (IYCF) [7, 20], an internationally recognized complementary feeding recommendation, to measure dietary diversity. The 2014 CDHS survey gathered data on types of food that children consumed the previous day. We categorized these food items into seven major foods groups based on the WHO’s IYCF guidelines such as (grains, roots, and tubers); (legumes and nuts); (flesh foods (meat, fish, poultry, and liver/organ meats)); (eggs); (vitamin A-rich fruits and vegetables); (dairy products (milk, yogurt, cheese)); and (Other fruits and vegetables). A food group was received a value of one (1) for a child if they consumed at least one foods item from it the previous day, and a value of zero (0) if they didn’t. The dietary diversity score, which ranges from zero to seven, whereby zero represents the non-consumption of any of the food items in the food groups, and seven represents the highest level of diet diversification. A child had reached the MDD if a child had consumed at least four of the seven food groups over the previous day.

#### Minimum meal frequency (MMF)

The minimum number of times the child consumed solid, semi-solid or soft foods within the prior 24□hours, day or night before the survey (including two milk feeds for non-breastfed children). The minimum frequency is twice for breastfed infants aged 6 to 8 months, three times for infants aged 9 to 23 months, and four times for non-breastfeed infants 6-23 months [21].

minimum meal frequency and diversified diet. A composite indicator of minimum dietary diversity and minimum meal frequency is the **minimum acceptable diet (MAD)**. Proportion of children aged 6-23 months who had a minimum variety of foods and a minimum meal frequency (other than breast milk) within 24□hours day or night before the survey [21].

#### Socio-demographics of study population

Children’s age in months were (6-8, 9-11,12-17, and 18-23), alone with their genders and whether they breastfed (yes vs no). Mother was divided into three age groups: 15-19, 20-34, and 35-49. Education level (no education, primary and secondary and higher). Employment of mothers (working vs not working). Residency (urban vs rural). Household wealth status (poor, middle, and rich). Piped into the home, piped into the yard or plot, public taps or standpipes, tubed wells or boreholes, protected wells, protected springs, and rainwater were all examples of improved sources of drinking water. The classification of other sources as non-improved. A toilet facility is one that has a flush toilet that is connected to a sewage or septic tank as well as a ventilated or improved latrine or other types of toilets. Facilities without toilets are classified as non-improved sanitation facilities.

### Statistical analysis

STATA version 16 (Stata Corp 2019, College Station, TX) was used for data cleaning and analysis. The STATA command “survey” package was used to declare that complex survey design was accounted for, and the standard sampling weight (v005/1,000,000), clustering, and stratification variables that were provided by DHS were used to perform all estimations by using survey-specific command “svy” [22, 23].

The results presented using *p*-values, and a bivariate Pearson chi-square test was used to assess significant associations between independent variables of interest such as children characteristics, the characteristics of mother, and household characteristics, geographical regions, and the feeding practice consumed and wasting status. A covariate was considered significant if it had a p-value <u>less than</u> 0.15 and was included in the analysis of multiple logistic regression [24]. Based on the literature that was examined and found to have an association with undernutrition, some potential confounding factors were included, such as children age, water drinking and feeding practices consumption [17, 18].

The magnitude effect of unadjusted associations between wasting and children, maternal, and household characteristics, geographical regions, and feeding practice consumption was examined using simple logistic regression. Odds Ratios (OR) with 95% confidence intervals (CI) and P-value are used to present the results. After adjusting for other potential confounding factors in the model, multiple logistic regression analysis was used to investigate the primary risk factors associated linked to wasted. The final multivariate model’s finding is reported as adjusted odds ratios (AOR) with 95% confidence intervals and corresponding p-values. Based on the final multivariable logistic regression model’s results having p-value less than 0.05 and 95% confidence intervals, the finding was considered as statistically significant.

### Ethics statement

The CDHS 2014 was approved by the Cambodia National Ethics Committee for Health Research (Ref: # 056 NECHR) and the Institutional Review Board (IRB) of ICF in Rockville, Maryland, USA. The CDHS data are publicly accessible and were made available to the researchers upon request to the DHS Program, ICF at https://www.dhsprogram.com.

## RESULTS

### Describes of study population characteristics

Describes the socio-demographic characteristics, mother, and household of 1,415 children aged 6-23 months in the survey. The mean age of children was 14.5 (+5.2 months), 35% were aged 18-23 months, and 17.2% were aged 6-8 months respectively. More than half (52%) of children were boys. Slightly more than 68.4% of children are currently breastfeed. Average age of mother was 27.5 (SD=5.9 years). Majority (82.5%) of them were aged 20-34 years, and 5.5% were aged 15-19 years. 12.6% of the mothers had no formal education, while 55.3% had completed their primary education. However, 31.7% of them were unemployed or not working. Many of the children 86.5% were born in rural areas. Slightly more than 44.8% of households were considered poor based on wealth index quintiles. Close to 52.6% of households had improved drinking water, and half of them had improved or not shared sanitation facilities. For wasting, prevalence was found to be at 10.4% (**Table 1**)

**Table 1.** Characteristics of the children aged 6-23 years old (n=1,415 weighted)

| **Variables** | **Freq.** | **%** |
| --- | --- | --- |
| **Age of children, mean(±SD) (months)** | 14.5(±5.2) | |
| 6-8 | 243 | 17.2 |
| 9-11 | 216 | 15.3 |
| 12-17 | 467 | 33.0 |
| 18-23 | 489 | **34.5** |
| **Sex of children** |  |  |
| Girls | 679 | 48.0 |
| Boys | 736 | **52.0** |
| **Breastfeeding** |  |  |
| No | 447 | 31.6 |
| Yes | 968 | **68.4** |
| **Mother's mean (±SD) age (years)** | 27.5(±5.9) | |
| 15-19 | 78 | 5.5 |
| 20-34 | 1167 | **82.5** |
| 35-49 | 170 | 12.0 |
| **Maternal educational** |  |  |
| No education | 179 | **12.6** |
| Primary | 782 | 55.3 |
| Secondary and higher | 454 | 32.1 |
| **Maternal working status (n=1,414)** |  |  |
| Yes | 838 | 59.3 |
| No | 576 | **40.7** |
| **Place of residence** |  |  |
| Rural | 1224 | **86.5** |
| Urban | 191 | 13.5 |
| **Household wealth status** |  |  |
| Rich | 503 | 35.6 |
| Middle | 278 | 19.6 |
| Poor | 634 | **44.8** |
| **Drinking water source in dry season** |  |  |
| Improved | 744 | 52.6 |
| Unimproved | 671 | **47.4** |
| **Toilet facility** |  |  |
| Improved | 704 | 49.7 |
| Unimproved | 711 | **50.3** |
| **Prevalence of Wasting** |  |  |
| Wasting | 1268 | **89.6** |
| Not wasting | 148 | **10.4** |

### Distributions of child feeding practices

The prevalence of child aged 6-23 months, received an minimum meal frequency at 71%, 71.6% for breastfed and 28.4% for non-breastfed. And met the minimum dietary diversity were 46.7%, 58.2% for breastfed and 41.8% for non-breastfed. Moverover, adequate minimum acceptable diet were 31.2%, 72.2% for breastfed and 27.8% for non-breastfed (**Fig 2**).

**Figure 2.**
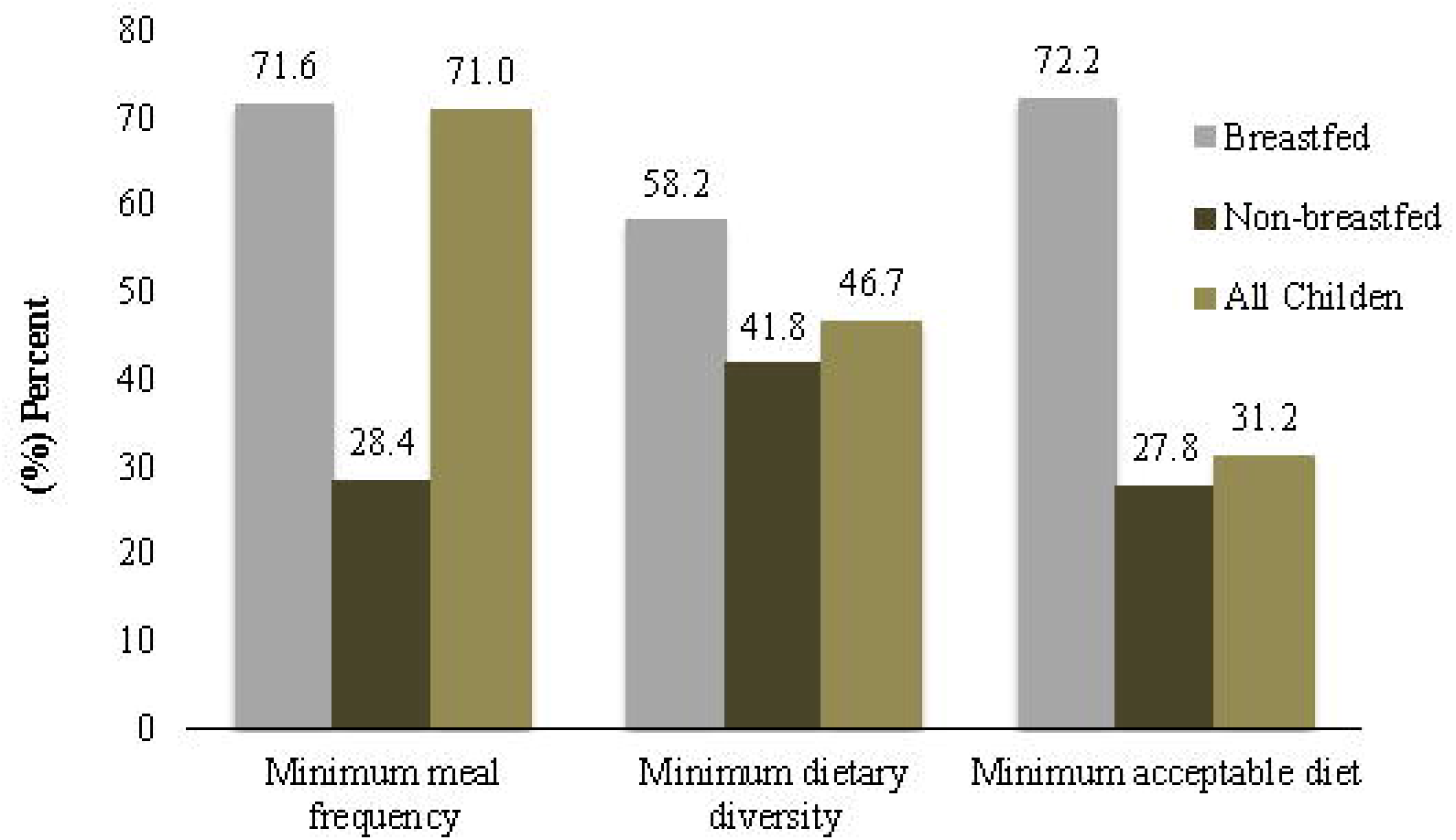
Indicators of complementary feeding practice of children 6-23 months of age in Cambodia, in 2014 (n=1,415)

### Association between feeding practice consumption and socio-demographics and wasting

As **table 2** shown, children who had inadequate dietary diversity was found 10% of children wasting, (p value =0.758). Inadequate meal frequency was 10.6% of wasting, with (p value =0.796). And unaccepted diet was 10.6% of wasting, with (p value =0.939), compared to those adequate. Similar, children aged 9-11 months was 13.5% of wasting compared to another age group with, (p value=0.370). Begin girls was 12% of wasting, compared to boys’ children with, (p value=0.217). None-breastfeeding practice children was 11.1% of wasting, compared to breastfeeding with, (p value=0.342). Children born from mothers were aged (15-19) were more likely wasting (25.9%) than older mothers, (p value= **0.001)**. While their mother had limited education had high proportion of wasting (16.3%), compared to educated, with (p value=0.179). Children born in rural areas were found to be more wasted (11%), compared to those in urban areas, with (p value=**0.077**). Children who live with poor household based on wealth index quintiles was found to be more wasted (14%), compared to those better wealth index quintiles, with (p value=**0.018)**. Moreover, children who from unimproved water, toilet facility source recorded higher prevalence of wasting was 12.3%, and 13.1% compared to those who from an improved source, with (p value=**0.015)**.

**Table 2.**
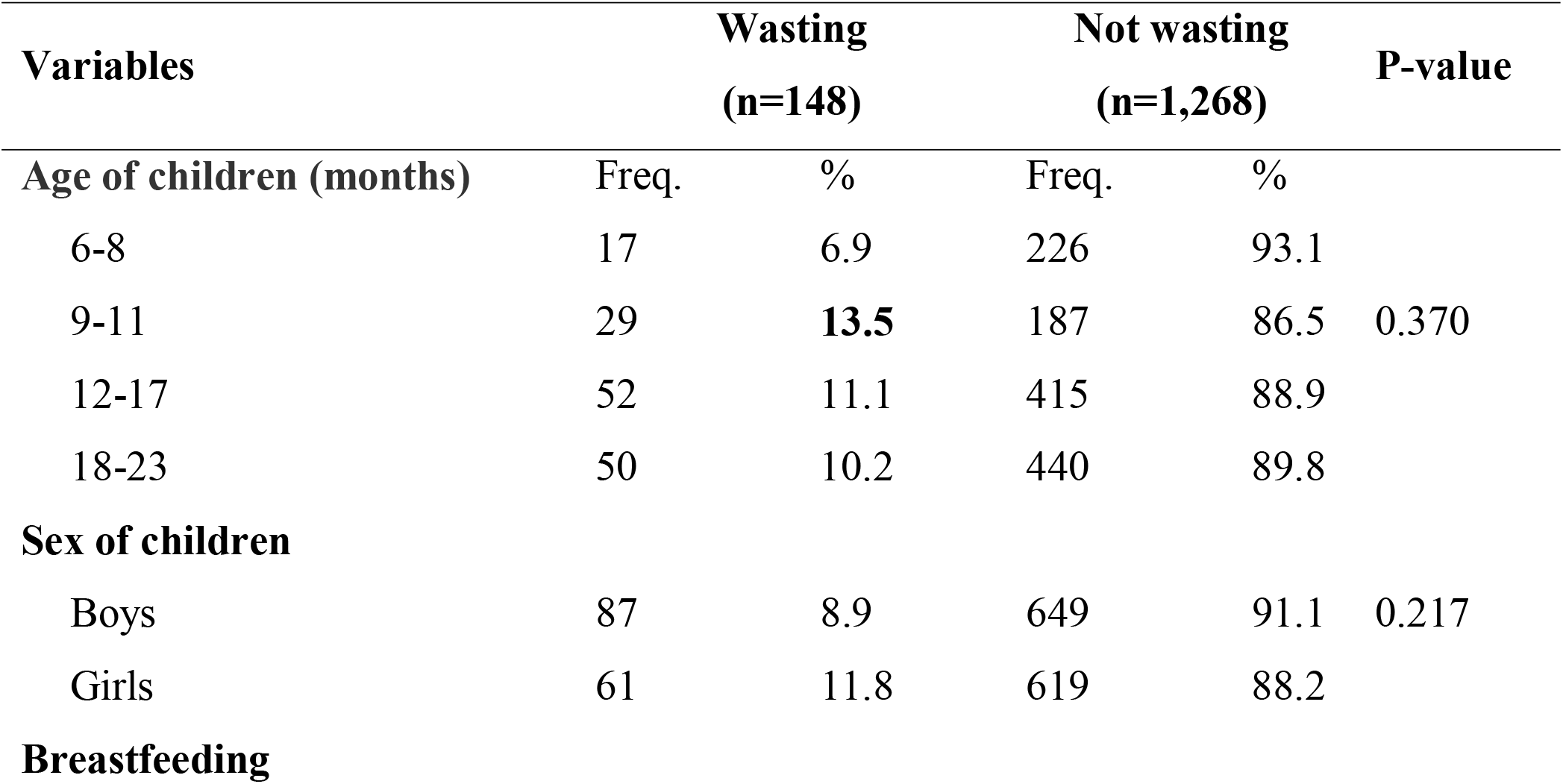
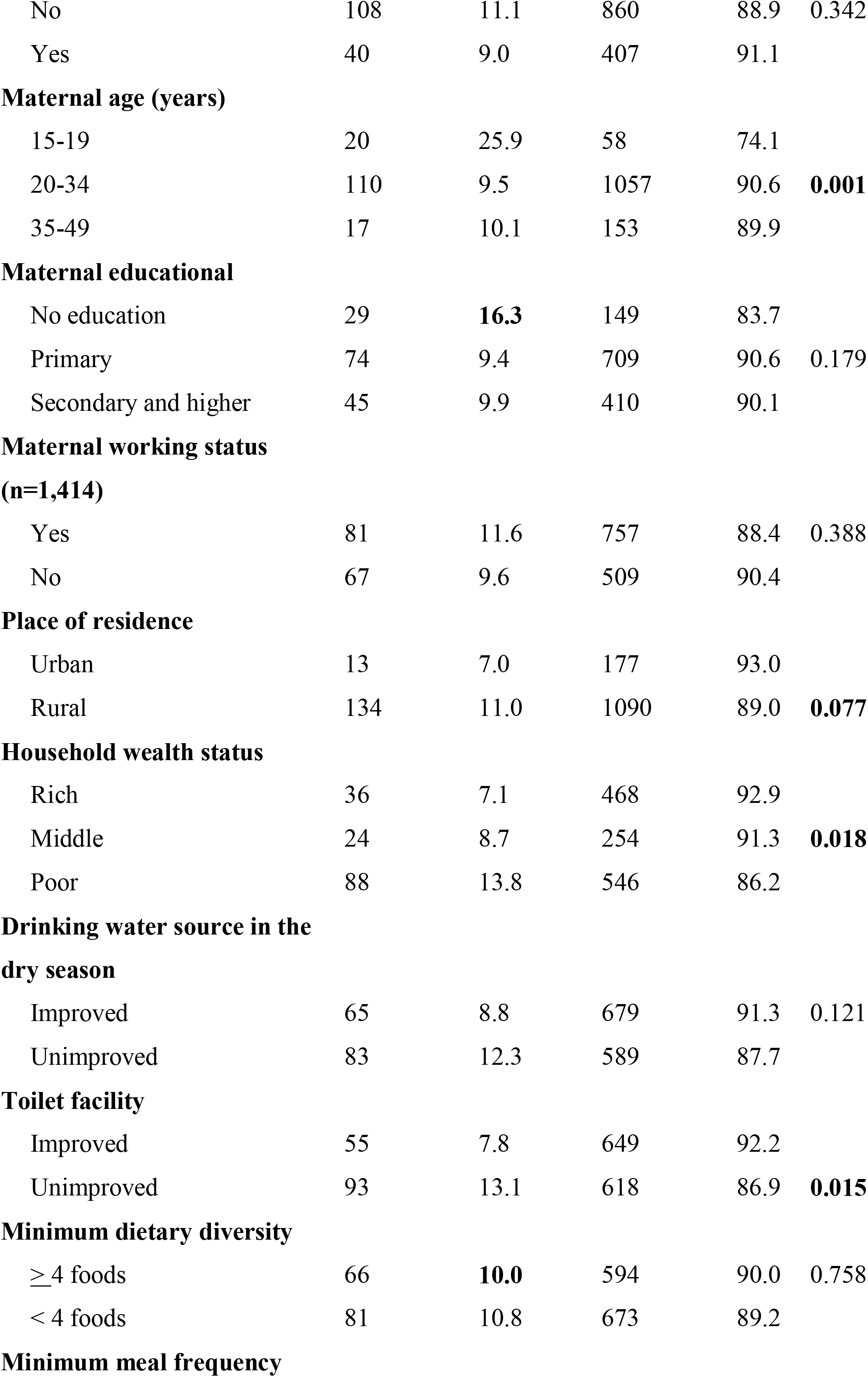
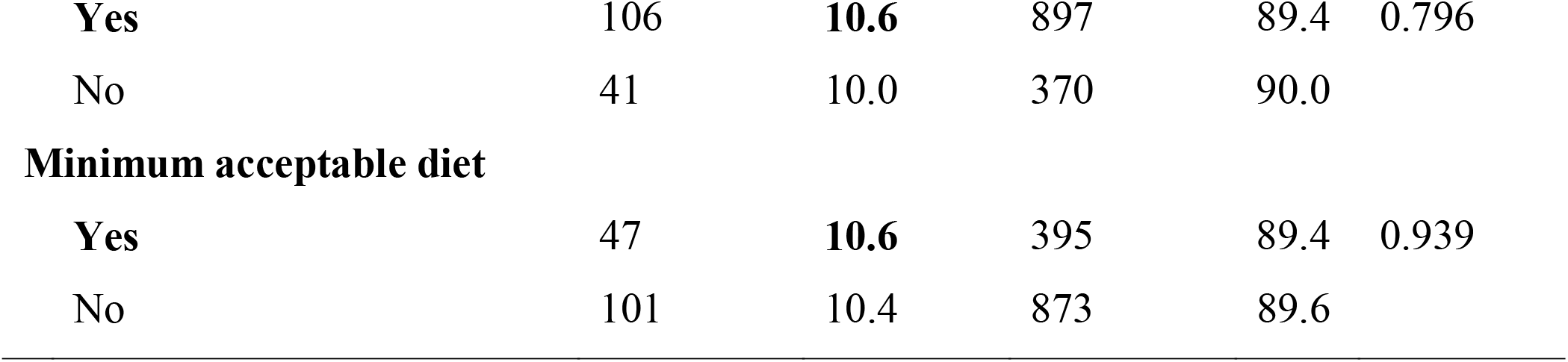
Bivariate analysis of the association between socio-demographic, dietary diversity, meal frequency, acceptable diet, and wasting (n=1,415 weighted)

### Multiple logistic regression results between feeding practice consumption and socio-demographics and wasting

Several factors were independently associated with wasting. Those children who aged 9-11 months were 2.3 times more likely to be wasted [AOR=2.3; 95% CI 1.0-5.0] compared to children aged 6-8 months, respectively. However, children whose mothers were in the aged 20-34 years and 35-49 years presented 70% reduced odds of wasting [AOR=0.3; 95% CI 0.2-0.6], and [AOR=0.3; 95% CI 0.1-0.7], compared to mother aged 15-19 years. Lastly, children from poor households were twice time more like to be wasted [AOR=1.9; 95% CI 1.1-3.5] compared to better household (**Table 3**).

**Table 3.** Univariate and multiple logistic regression analysis of the association between food groups, minimum dietary diversity, and wasting

| Variables | (n=1,415 weighted) |  |  | (n=1,415 weighted) |  |  |
| --- | --- | --- | --- | --- | --- | --- |
|  | OR | 95% CI | P-value | AOR | 95% CI | P-value |
| **Age of children (month)** |  |  |  |  |  |  |
| 6-8 | 1.0 | Ref. |  | 1.0 | Ref. |  |
| 9-11 | 2.1 | 1.0-4.5 | 0.055 | **2.3** | **1.0-5.0** | **0.036** |
| 12-17 | 1.7 | 0.8-3.5 | 0.172 | 1.7 | 0.8-3.7 | 0.164 |
| 18-23 | 1.5 | 0.8-3.0 | 0.230 | 1.8 | 0.9-3.8 | 0.093 |
| **Sex of children** |  |  |  |  |  |  |
| Boys | 1.0 | Ref. |  | - | - |  |
| Girls | 0.7 | 0.4-1.2 | 0.219 | - | - |  |
| **Breastfeeding** |  |  |  |  |  |  |
| No | 1.0 | Ref. |  | 1.0 | Ref. |  |
| Yes | 1.3 | 0.8-2.1 | 0.343 | 1.2 | 0.7-2.3 | 0.518 |
| **Maternal age (years)** |  |  |  |  |  |  |
| 15-19 | 1.0 | Ref. |  | 1.0 | Ref. |  |
| 20-34 | 1.5 | 0.7-3.4 | 0.274 | **0.3** | **0.2-0.6** | **0.001** |
| 35-49 | 2.4 | 1.0-5.9 | 0.052 | **0.3** | **0.1-0.7** | **0.007** |
| **Maternal educational** |  |  |  |  |  |  |
| No education | 1.0 | Ref. |  | - | - |  |
| Primary | 0.9 | 0.5-1.4 | 0.611 | - | - |  |
| Secondary and higher | 0.7 | 0.4-1.1 | 0.121 | - | - |  |
| **Maternal working status** |  |  |  |  |  |  |
| Yes | 1.0 | Ref. |  | - | - |  |
| No | 0.8 | 0.6-1.1 | 0.109 | - | - |  |
| **Place of residence** |  |  |  |  |  |  |
| Urban | 1.0 | Ref. |  | 1.0 | Ref. |  |
| Rural | 1.5 | 1.1-2.2 | 0.024 | 1.1 | 0.6-2.1 | 0.785 |
| **Household wealth status** |  |  |  |  |  |  |
| Rich | 1.0 | Ref. |  | 1.0 | Ref. |  |
| Middle | 1.8 | 1.1-2.9 | 0.020 | 1.2 | 0.6-2.7 | 0.626 |
| Poor | 2.5 | 1.7-3.5 | 0.000 | **1.9** | **1.1-3.5** | **0.034** |
| **Drinking water source in dry season** |  |  |  |  |  |  |
| Improved | 1.0 | Ref. |  | 1.0 | Ref. |  |
| Unimproved | 1.4 | 1.1-2.0 | 0.022 | 1.4 | 0.8-2.3 | 0.215 |
| **Toilet facility** |  |  |  |  |  |  |
| Improved | 1.0 | Ref. |  | - | - |  |
| Unimproved | 2.2 | 1.6-3.1 | 0.001 | - | - |  |
| **Minimum dietary diversity** |  |  |  |  |  |  |
| $\geq 4$ foods | 1.0 | Ref. | | 1.0 | Ref. | |
| $< 4$ foods | 1.1 | 0.7-1.8 | 0.758 | 1.2 | 0.6-2.2 | 0.626 |
| **Minimum meal frequency** |  |  |  |  |  |  |
| **Yes** | 1.0 | Ref. |  | 1.0 | Ref. |  |
| No | 0.9 | 0.6-1.5 | 0.796 | 0.9 | 0.5-1.4 | 0.556 |
| **Minimum acceptable diet** |  |  |  |  |  |  |
| **Yes** | 1.0 | Ref. |  | 1.0 | Ref. |  |
| No | 1.0 | 0.6-1.6 | 0.939 | 0.8 | 0.4-1.6 | 0.472 |

## DISCUSSION

The association between child feeding practices consumption factors like dietary diversity, meal frequency, acceptable diet and child wasting was not discovered in this study. In line with published studies [25, 26]. In contract to feeding practices consumption, wasting is associated with maternal younger age, child age, childhood diseases and lowest household wealth qualities rather than [27]. This could be because acute malnutrition children that results of shorter-term episodes of inadequate feeding or illnesses is referred to as “wasting” [27]. However, eating a variety of food plays a significant part in reducing wasting among young children. According to findings from other studies conducted in Sub-Saharan Africa [26], South Ethiopia [28], and Burundi and Democratic Republic Congo [29], children who had an adequate minimum dietary diversity had lower odds of wasting than those who had an inadequate minimum dietary diversity. Meaning they consume at least four of the seven foods groups-grains, roots, and tubers; legumes and nuts; dairy products; flesh foods (meat, fish, poultry, and organ meats); eggs; vitamin A rich fruits and vegetables; other fruits and vegetables-at a time [30]. Furthermore, we found several main predictors of child wasting in the study included children aged 9-11 months and children from poor household wealth status having higher odds of being wasting. However, mothers aged 20-49 years old had decreased odds of having wasting. Children from poorest households high prevalence of wasting, according to a study in 35 low income and middle income countries [31]. According to research conducted in South East Asia, children from the poor wealth quintiles were 25% more likely to be wasted than compared to the richest wealth quintile [32]. Due to their limited access to sufficient food of quality and their substandard living conditions, households with low incomes tend to spend less on proper nutrition and are more susceptible to growth failure.

Our study found children who born from older mother age (20-49 years) were protected about 60% of being wasting, compared to who born from young mother. In line with study conducted in Tamale Metropolis, Ghana [33]. Possible explanations young mother may not have adequate financial resources to provide the needs of the babies owing to their poor socio-economic circumstances and that of their partners and their biological development for having babies.

The study has the number of limitations. First, the CDHS was a cross-sectional study, it was not possible to simultaneously assess the temporal associations, or causality between child feeding practices and wasting at the same times. Second, since mother and other caregivers self-reported the study’s findings, recall and measurement bias may have led to an overestimation of the food groups that children consumed over the course of a during 24 hours period. Thirdly, only a few maternal and child factors considered in this study, such as maternal diseases and childhood illness of diarrhea and fever and did not include other diseases. Other diseases were not considered. Also excluded from the analysis were variables such as mode of birth, place of birth, birth weight child and whether the child had received vaccinations. Furthermore, this which could did not account for total calories because to lack of data on calorie consumption from the CDHS and this could have impact on the extrapolations made from it. Finally, since these data were gathered in 2014, they might no longer accurately depict Cambodia’s current child feeding practices. When the data are made available to public, more analysis of the prevalence of wasting and its relationship to child feeding practices could be done. Despite the limitations, the study has some advantages. A key strength of the study is use of nationally representative data and a large sample size. Additionally, the findings help fill a gap in literature on Cambodia’s varied diet and wasting.

In conclusion, the results of this study demonstrate that wasting is common problem among Cambodian children aged 6-23 months, and a fewer than half of them consumed the recommended minimum amount of dietary diversity. Despite not being discovered in this study, it was found in other studies. Therefore, it is advised that interventions and policy development should place a high priority on encouraging children in Cambodia to consume a variety of foods.

Additionally, children aged 6-11 months and children of young mothers should receive special attention from the intervention to reduce wasting. Designing, children’s nutrition program interventions with focus on these socio-demographic determinants it therefore essential. A possible intervention to increase food diversity consumption and to decrease child waste is to improve knowledge and practices regarding the importance of feeding children a diverse diet, as food consumption for children aged 6-23 months is primarily dependent on caregivers. The two best complementary feeding practices and strengthening exclusive breastfeeding for at least two years could be the potential interventions to reduce child wasting in Cambodia.

## Data Availability

All data produced in the present study are available upon reasonable request to the authors

https://www.dhsprogram.com.

## ACKNOWLEDGMENTS

The authors would like to thank DHS-ICF, who approved the data used for this paper.

## AUTHORS’ CONTRIBUTIONS

**Um S**. contributed to conceptualization, methods, data analysis, and writing original draft, writing review & editing; **Mom L**. contributed to conceptualization, methods, data analysis, and writing original draft, writing review & editing; **An Y**. contributed to writing review & editing manuscript. **Bunkea T**. contributed to writing review & editing manuscript. All authors read and approved the final manuscript.

## DATA AVAILABILITY

Our study used the 2014 Cambodia Demographic and Health Survey (CDHS) datasets. The DHS data are publicly available from the website: https://www.dhsprogram.com/data/dataset_admin

## ABBREVIATIONS

WHO: World Health Organization
CDHS: Cambodia Demographic Health Survey
U5: Children Under Five
MMD: Minimum Dietary diversity
MMF: Minimum meal frequency
MAD: Minimum acceptable diet
AOR: Adjusted odds ratio
PPS: Probability proportional to size
EA: Enumeration areas.

